# Efgartigimod efficacy and safety in refractory Myasthenia Gravis - UK’s first real-world experience

**DOI:** 10.1101/2024.01.31.24302082

**Authors:** J Moniz Dionísio, P Ambrose, G Burke, M Farrugia, P Garcia-Reitboeck, C Hewamadduma, M Hill, RS Howard, S Jacob, DM Kullmann, MI Leite, J Miller, A Pinto, J Pritchard, T Riswick, S Sathasivam, N Thambarigjah, S Viegas, F Norwood, J Spillane

**Author notes:** This study did not receive any funding.

## Abstract

**Background:** We report our experience of patients with generalised MG (gMG) treated with Efgartigimod, an FcN antagonist, under the Early Access to Medicine Scheme (EAMS) in the UK.

**Methods:** Data from all UK patients treated with Efgartigimod under the EAMS June 22-July 23 were collected retrospectively. Efgartigimod was administered as per the ADAPT protocol (consisting of a treatment cycle of 4 infusions at weekly intervals with further cycles given according to clinical need).

**Results:** 48 patients with AChR antibody-positive gMG were treated in 12 centres. Most (75%) were female and most had a disease duration of over 10 years. The average MG-ADL score at baseline was 11.2. Most (72.9%) patients had undergone thymectomy. 77.0% were taking prednisolone at baseline. All patients had utilized non-steroidal immunosuppressant treatments, the average number tried was 2.6 (range 1-6). 51% had received Rituximab. 54.2% of patients required regular IVIg/PLEX.

75% of patients had a mean reduction in the MG-ADL of ≥2 points in the first cycle and this remained stable throughout the study. The mean intracycle reduction in the MG-ADL score in the 1st, 2nd, 3rd and 4th cycles were -4.6, -3.9, -3.4 and -4.2 respectively. Side effects were generally mild though one patient stopped treatment due to severe hypokalemia. No rescue treatments were required. At the end of the study, 96% of patients remained on Efgartigimod.

**Conclusion:** Efgartigimod is a safe and effective treatment for patients with refractory, treatment-resistant gMG.

## INTRODUCTION

Myasthenia Gravis (MG) is an autoimmune disorder of the neuromuscular junction that causes fatigable neuromuscular weakness. 85% of patients have antibodies against the acetylcholine receptor (AChR) and a varying proportion of the remainder have antibodies against muscle-specific kinase (MuSK), an important post-synaptic clustering protein. A smaller proportion of patients have antibodies against low-density lipoprotein receptor-related protein 4 (LRP-4), another post-synaptic protein. Up to 10% of patients do not have detectable antibodies on the conventional assay but about one-third of this cohort will have antibodies detectable on a more specific cell-based assay^1^.

The hallmark of MG is fatigable neuromuscular weakness of skeletal muscles. Ocular symptoms – diplopia and ptosis can occur alone, but most patients develop generalized symptoms of fatigable limb weakness, facial weakness, and difficulties with speech, chewing and swallowing. Up to 15% of patients can develop myasthenic crisis; or ventilatory failure due to respiratory muscle weakness^2^.

MG is a treatable neuromuscular disease and due to advances in the care of patients with MG including progress in intensive care, there has been a reduction in the mortality of MG from 70% in the 1930s to <10% nowadays^3^. The first line treatment is pyridostigmine which can provide short-term symptomatic relief but has no disease-modifying effect. Surgical removal of the thymus is almost always indicated for patients with a thymoma, but MG symptoms will generally persist post-operatively. In patients without a thymoma, thymectomy has been shown in a randomized controlled trial to be associated with better outcomes in younger onset seropositive generalized MG but the effects are variable and may not be evident for many years^4^. Treatments such as intravenous immunoglobulin (IVIg) and therapeutic plasma exchange (TPE) can improve symptoms rapidly, though the effects are short-lasting, and these treatments are generally reserved for acute severe exacerbations. The mainstay of management of MG therefore rests on nonspecific broad-spectrum immunosuppression with steroids and non-steroid immunosuppressant therapies such as azathioprine, mycophenolate, and methotrexate, with Rituximab occasionally used in refractory patients who demonstrate active disease despite treatment with at least two trials of immunosuppressant agents^5^.

Despite advances in treatment, there is a clear unmet need for patients with MG^5^. Steroids though effective are associated with a plethora of well-documented side effects. Immunosuppressive agents are not tolerated in many patients with one review showing that over 30% of patients taking azathioprine have adverse events and up to 11% have to discontinue the drug^6,7^. Moreover, oral immunosuppressants have a slow onset of action meaning that patients are exposed to high-dose steroids for months and sometimes years. Approximately 15% of patients are refractory to standard therapies and are often dependent on costly treatments such as IVIg and TPE^3^. Real-world studies have shown that over 40% of patients with MG have unacceptable disease control^8^ and MG is known to have a significant impact on quality of life^9^.

There has therefore been a longstanding need for more targeted therapies in MG that are more efficacious and have a faster onset of action with a favorable side-effect profile. Advances in the understanding of the pathogenesis of MG have unveiled several potential treatment targets including B cells, complement, IL-6 and other cytokines^2^.

A novel therapeutic target that has emerged in recent years is the neonatal FC receptor (FcRN). This is a ubiquitous MHC Class 1-like molecule that allows the recycling of IgG by protecting it from lysosomal degradation thus prolonging its half-life. Efgartigimod (ARGX-113) is a human IgG Fab fragment that has been engineered to have a higher affinity for the Fc receptor than native IgG, thus reducing IgG recycling and lowering IgG levels in circulation. The mechanism of lowering IgG levels can be thought of as analogous to therapeutic plasma exchange which is known to improve symptoms in MG rapidly but the availability of TPE is limited and can be associated with significant side effects.

In phase 1 and 2 trials Efgartigimod was found to significantly reduce IgG levels and improve symptoms in generalized MG^10,11^. The efficacy and safety of Efgartigimod were then studied in a phase 3 double-blind randomized placebo clinical trial (the ADAPT trial)^12^. Efgartigimod was given as an intravenous (IV) infusion weekly for four weeks with further cycles repeated as necessary (based on clinical progression) no sooner than 8 weeks after the beginning of the previous cycle. Efgartigimod was found to be safe and well tolerated and a significantly higher number of patients treated with Efgartigimod met the primary outcome: the number of patients who had more than a 2-point reduction in the MG Activities of Daily Living (MG-ADL) score sustained for >4 weeks in the first treatment cycle. The MG-ADL score is a patient-reported outcome measure that allows an estimation of MG symptoms and their impact on daily living activities, ranging from 0 to 24, with greater values related to severe symptoms^13^.

Efgartigimod alfa is now approved by the FDA and the EMA. It was licensed by the Medicines and Healthcare Products Regulatory Authority (MHRA) in the United Kingdom (UK) in March 2023 but is not yet commercially available. However, it has been available for adults with generalised AChR antibody under the (MHRA) Early Access to Medicines Scheme (EAMS) in specialist UK MG centres since May 2022. The aim of an EAMS scheme is to provide earlier access to unlicensed treatments for patients with a high unmet clinical need. After licensing, Efgartigimod was made available under the EAMS Plus Scheme (EAMS +) whilst NICE approval was sought.

Under the EAMS scheme, Efgartigimod was indicated for the treatment of adult patients with AChR antibody-positive gMG including those who had failed, did not tolerate or were ineligible for licensed treatment. Patients could not have received Rituximab within 6 months or IVIg within 4 weeks and IgG levels had to be ≥ 6g/L prior to starting Efgartigimod. The consensus achieved before the introduction of the scheme with UK MG clinicians was that it would be reserved for patients with refractory disease who had not responded to ≥ 2 non steroids immunosuppressant agents who were intolerant or ineligible for such therapies and those patients who were dependent on IVIG and TPE. Clinicians also agreed to collect data regarding ADL scores, treatment failures and any adverse events.

According to the EAMS treatment protocol, Efgartigimod was given as per the ADAPT trial as a cyclical treatment as an IV infusion weekly for four weeks (10 mg/kg in one-hour intravenous infusion), with subsequent treatment cycles dependent on the patient’s symptoms. A home care service was provided for some patients after their initial cycle in hospital.

Our objective with this study was to provide the first real-world experience regarding the Efgartigimod efficacy, safety and tolerability in the UK population.

## METHODS

### Study design

This was a retrospective observational multicenter study designed to analyse the efficacy and safety of Efgartigimod in AChR antibody-positive generalised MG patients who met the EAMS criteria for its use.

### Participants

All UK MG specialist centres were invited to provide deidentified data regarding patients treated with Efgartigimod between March 2022 and July 2023.

### Outcomes

Our primary outcome was to determine the percentage of MG-ADL responders in the first cycle. Similar to what was defined in the ADAPT trial^11^, we considered that a patient had responded to the treatment when there was a 2-point reduction in the MG-ADL score sustained for >4 weeks. We also sought to understand the variation of MG-ADL at different time points (day 0, day 22 and day 36) for each cycle, to describe the incidence of adverse events and to determine the need for rescue treatments and the rate of Efgartigimod discontinuation.

### Statistical analysis

Statistical analysis was done using SPSS Statistics® version 28. Demographic data was presented descriptively, with mean values, standard deviation (SD), total number (N) and percentage (%). The variation of MG-ADL along the different time points, in each cycle, was analyzed with a mixed linear model for repeated measures, assuming missing at random data. A *p*-value of <0.05 was considered statistically significant.

STROBE cohort checklist was used when writing our report^14^.

## RESULTS

Our analysis included 48 patients from 13 centres who had completed at least one cycle of Efgartigimod under the EAMS scheme in the UK by 20^th^ July 2023. At the time, this represented 100% of MG patients who had completed at least one cycle of treatment. No patients were excluded from the analysis.

Most patients were female (75.0%, N = 36), with an average age of 49.2 (21.0 – 75.0, SD = 14.2) years old. The majority (66.7%, N = 32) had been diagnosed with MG more than 10 years before starting Efgartigimod. The average MG-ADL score at baseline was 11.2 (5 – 19, SD = 3.2). Most patients (72.9%, N = 35) had undergone thymectomy in the past (mean time since thymectomy = 12.5 years, 1 – 38, SD = 8.3).

All patients had utilized at least one non-steroidal immunosuppressant treatment (NSIST) in the past, and the average number tried prior to Efgartigimod was 2.6 (range 1 - 6). The most frequent NSISTs used included Azathioprine (79.2%, N = 38), Mycophenolate Mofetil (64.6%, N = 31) and Methotrexate (41.7%, N = 20). Six patients had received Cyclosporin, one had taken Tacrolimus and two had received Eculizumab. Just above a half (52.1%, N = 25) had previously received Rituximab.

70.8% (N = 34) had previously received IVIg and 43.8% (N = 21) were still requiring it on a regular basis at the time of Efgartigimod initiation. More than a quarter (27.0%, N = 13) had previously been treated with TPE in the previous year and 14.6% (N = 7) were still using it regularly at treatment initiation.

Just prior to the initiation of Efgartigimod, the majority of patients were taking a combination of NSIST and prednisolone (54.2%, N = 26). Ten patients were taking prednisolone only, and five were taking an NSIST only. Six patients were not on any immunosuppressive treatment at baseline though three of these patients were on regular IVIg. The NSISTs used included Azathioprine (7 patients), Mycophenolate Mofetil (14 patients), Methotrexate (8 patients) and Cyclosporin (2 patients). The average prednisolone dose was 20.5 mg daily (range 2-60 mg).

The reasons for starting Efgartigimod were listed as follows (participants could list more than one reason): refractory MG (77.0%, N = 37), the burden of treatment (35.4%, N = 17), dependent on IVIg/TPE (29.2%, N = 14), side-effects (12.5%, N = 6) and other reasons (4.2%, N = 2; participants specified needing bridging treatment). The detailed demographic and clinical data are available in Tables 1 and 2, respectively.

**Table 1.**
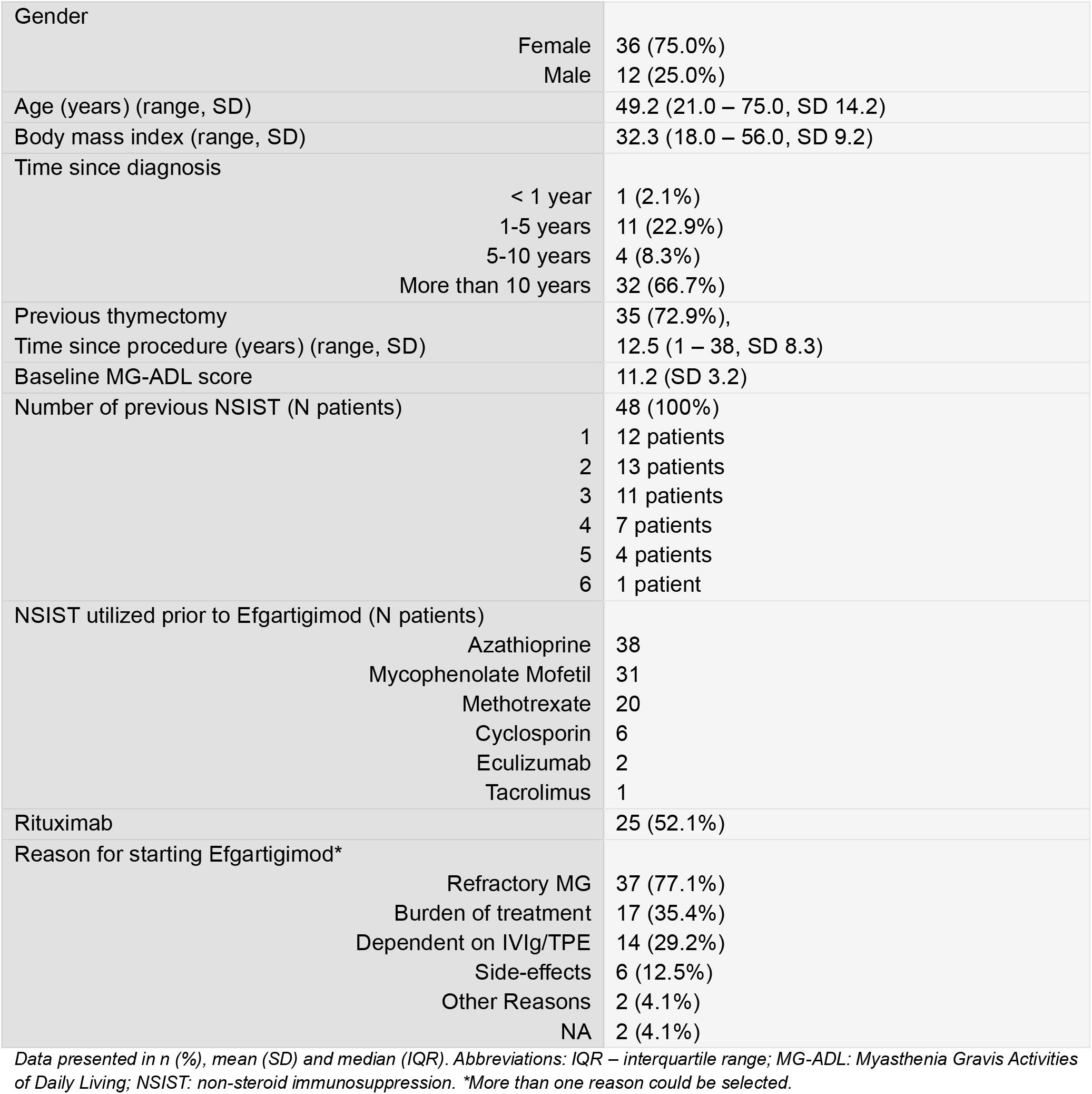
Demographic and Clinical Data (N = 48)

**Table 2.**
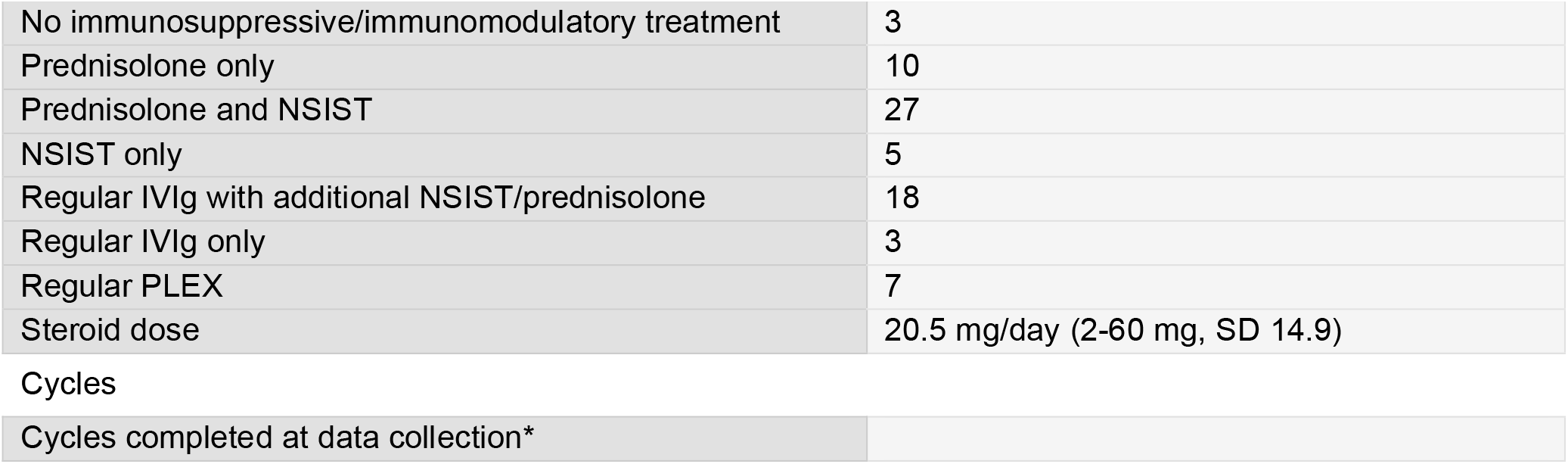
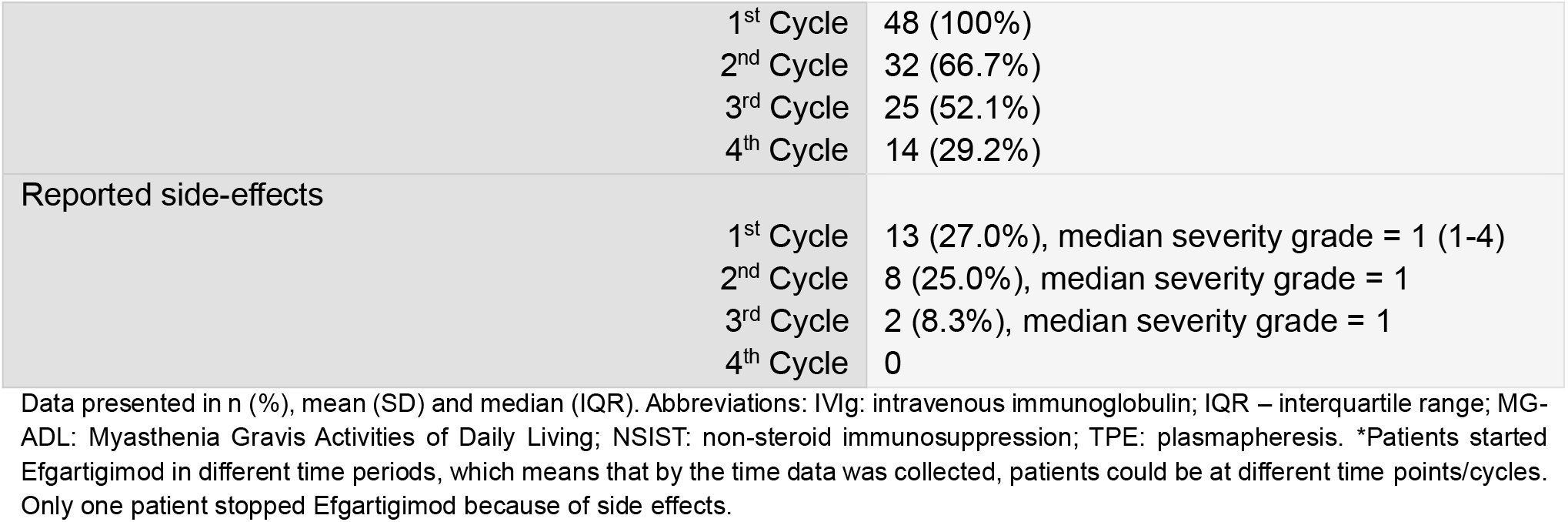
MG treatment at the time of commencing Efgartigimod and cycles completed (N = 48)

|  |  |
| --- | --- |
| No immunosuppressive/immunomodulatory treatment | 3 |
| Prednisolone only | 10 |
| Prednisolone and NSIST | 27 |
| NSIST only | 5 |
| Regular IVIg with additional NSIST/prednisolone | 18 |
| Regular IVIg only | 3 |
| Regular PLEX | 7 |
| Steroid dose | 20.5 mg/day (2-60 mg, SD 14.9) |
| Cycles |  |
| Cycles completed at data collection* |  |

|  |  |  |
| --- | --- | --- |
|  | 1<sup>st</sup> Cycle | 48 (100%) |
|  | 2<sup>nd</sup> Cycle | 32 (66.7%) |
|  | 3<sup>rd</sup> Cycle | 25 (52.1%) |
|  | 4<sup>th</sup> Cycle | 14 (29.2%) |
| Reported side-effects | 1<sup>st</sup> Cycle | 13 (27.0%), median severity grade = 1 (1-4) |
|  | 2<sup>nd</sup> Cycle | 8 (25.0%), median severity grade = 1 |
|  | 3<sup>rd</sup> Cycle | 2 (8.3%), median severity grade = 1 |
|  | 4<sup>th</sup> Cycle | 0 |

Data presented in n (%), mean (SD) and median (IQR). Abbreviations: IVIg: intravenous immunoglobulin; IQR – interquartile range; MG-ADL: Myasthenia Gravis Activities of Daily Living; NSIST: non-steroid immunosuppression; TPE: plasmapheresis. \*Patients started Efgartigimod in different time periods, which means that by the time data was collected, patients could be at different time points/cycles. Only one patient stopped Efgartigimod because of side effects.

In our population, 75.0% (36 patients, N = 48) were defined as responders in the first cycle as they achieved a reduction in the MG-ADL score of ≥ 2 points. This percentage decreased slightly in the following cycles but remained stable throughout the study: 65.6% were responders in the second cycle (21 patients, N = 32), 72.0% in the third cycle (18 patients, N = 25) and 64.3% in the last cycle (9 patients, N = 14). Four patients who were not responders on the first cycle responded to the second cycle.

The mean reduction in MG-ADL score at the end of each cycle (day 22) comparing to its beginning (day 0) was, respectively, -4.6 points in the 1^st^ cycle, -3.9 points in the 2^nd^ cycle, -3.4 points in the 3^rd^ cycle, and -4.2 in the 4^th^ cycle (see Figure 1). When comparing the MG-ADL score at the end of each cycle versus the beginning of treatment with Efgartigimod (see Table 3), the mean reductions were: -4.5 points for cycle 1, -6.0 points for cycle 2, -6.9 points for cycle 3 and -7.8 points for cycle 4 (considering just the patients that completed each cycle: 48, 32, 23 and 14).

**Table 3.**
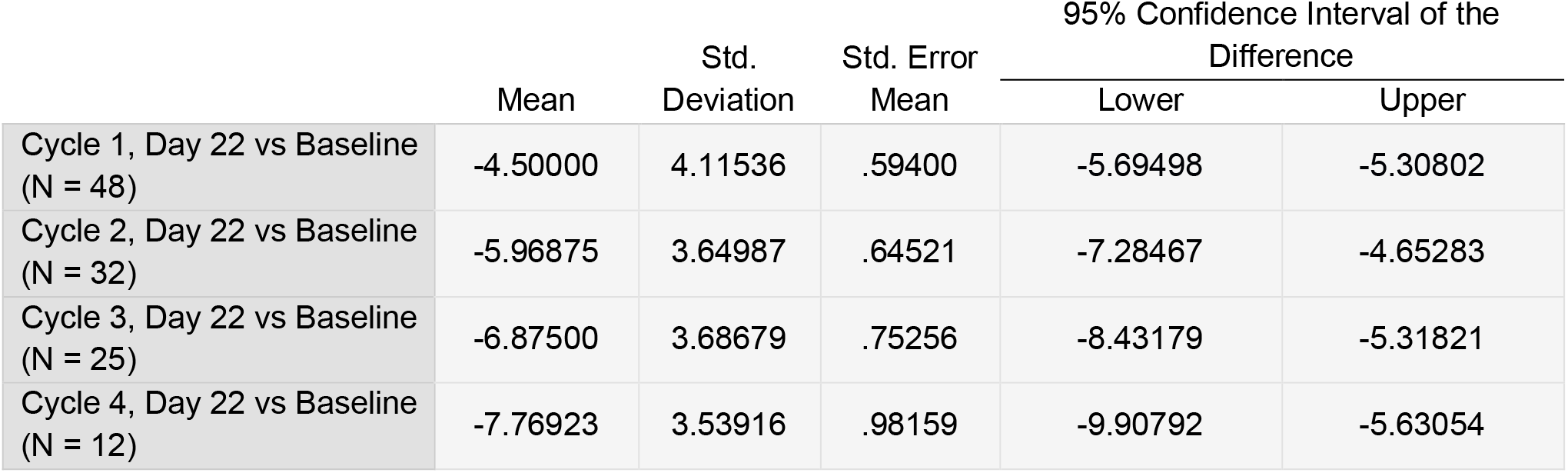
The mean difference in MG-ADL Scores comparing MG-ADL at baseline with the last day of each cycle.

**Figure 1.**
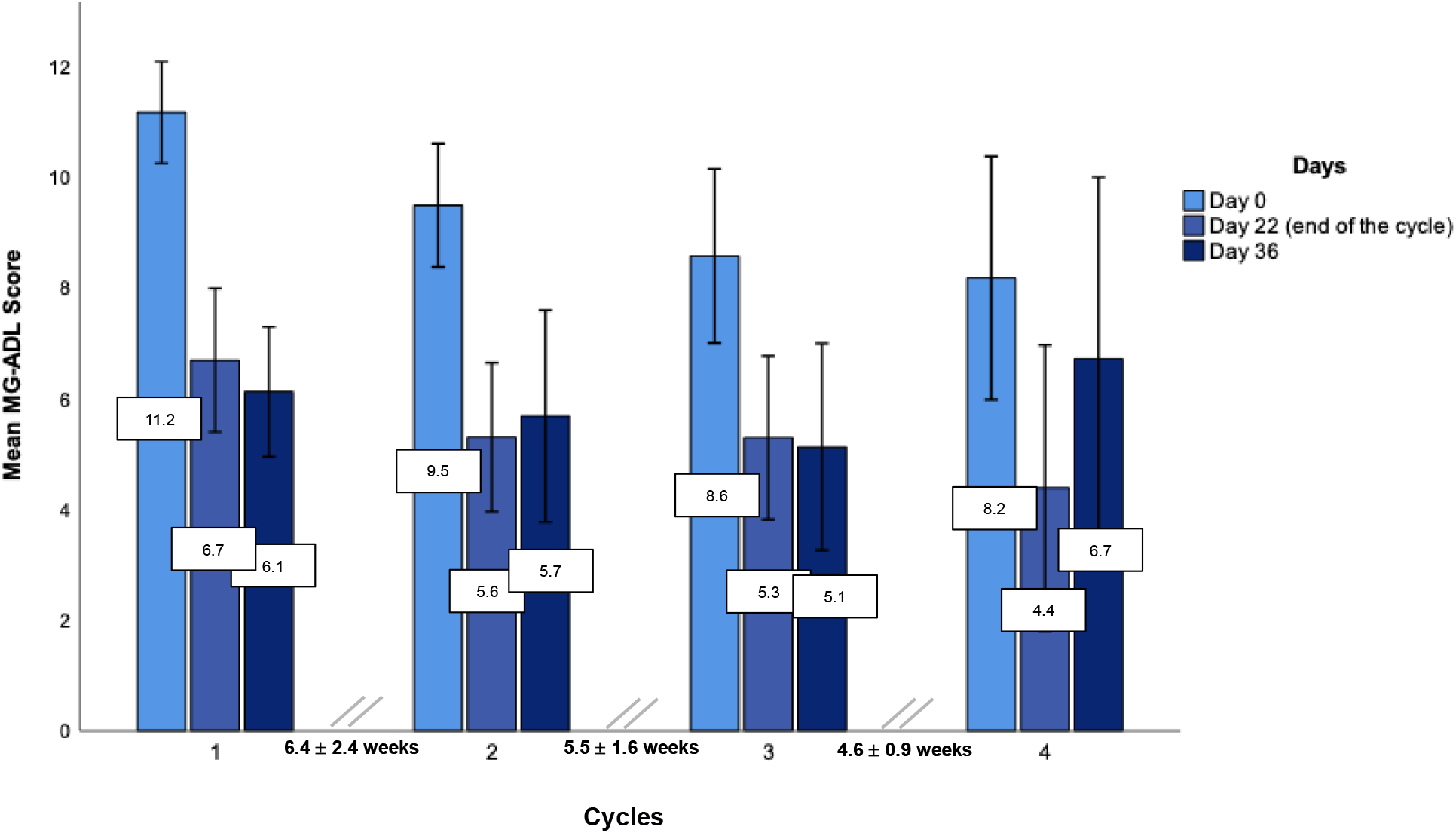
Mean intracycle MG-ADL score (Cycle 1, N = 48; Cycle 2, N = 32; Cycle 3, N = 23; Cycle 4, N = 14)

Using an MG-ADL score of 0 or 1 to define Minimal Manifestation Status, we observed that 10.4% (5 patients, N = 48) of all patients achieved this status by the end of the 1^st^ cycle. By the end of the 2^nd^ cycle, 12.5% (4 patients, N = 32) reached MMS; this was achieved by 14.3% by the end of the 3^rd^ cycle (4 patients, N = 25) and 35.7% by the end of the 4^th^ cycle (5 patients, N = 14).

A two-way repeated measures ANOVA with a Greenhouse-Geisser correction was performed in the subgroup that completed the first three cycles and analysed the longitudinal variation considering day 0 and day 22 (N = 25). The mean MG-ADL score differed statistically significantly between time points (p < 0.01). A *post-hoc* analysis with a Bonferroni adjustment confirmed that the MG-ADL score was statistically significantly decreased from pre-treatment to the end of the 1^st^ cycle (mean reduction of 5.54, p < 0.001, 95% CI, 3.26 – 7.82), to the end of the 2^nd^ cycle (mean reduction of 6.7, p < 0.001, 95% CI, 4.46 – 8.96), to the beginning of the 3^rd^ cycle (mean reduction of 3.1, p = 0.032, 95% CI, 0.17 – 6.08) and to the end of the 3^rd^ cycle (mean reduction of 6.9, p < 0.001, 95% CI, 4.4 – 9.3). The complete analysis between each cycle is shown in Figure 2 and the full analysis of the MG-ADL score variation in the three cycles can be found in the supplementary material (Table 1).

**Figure 2.**
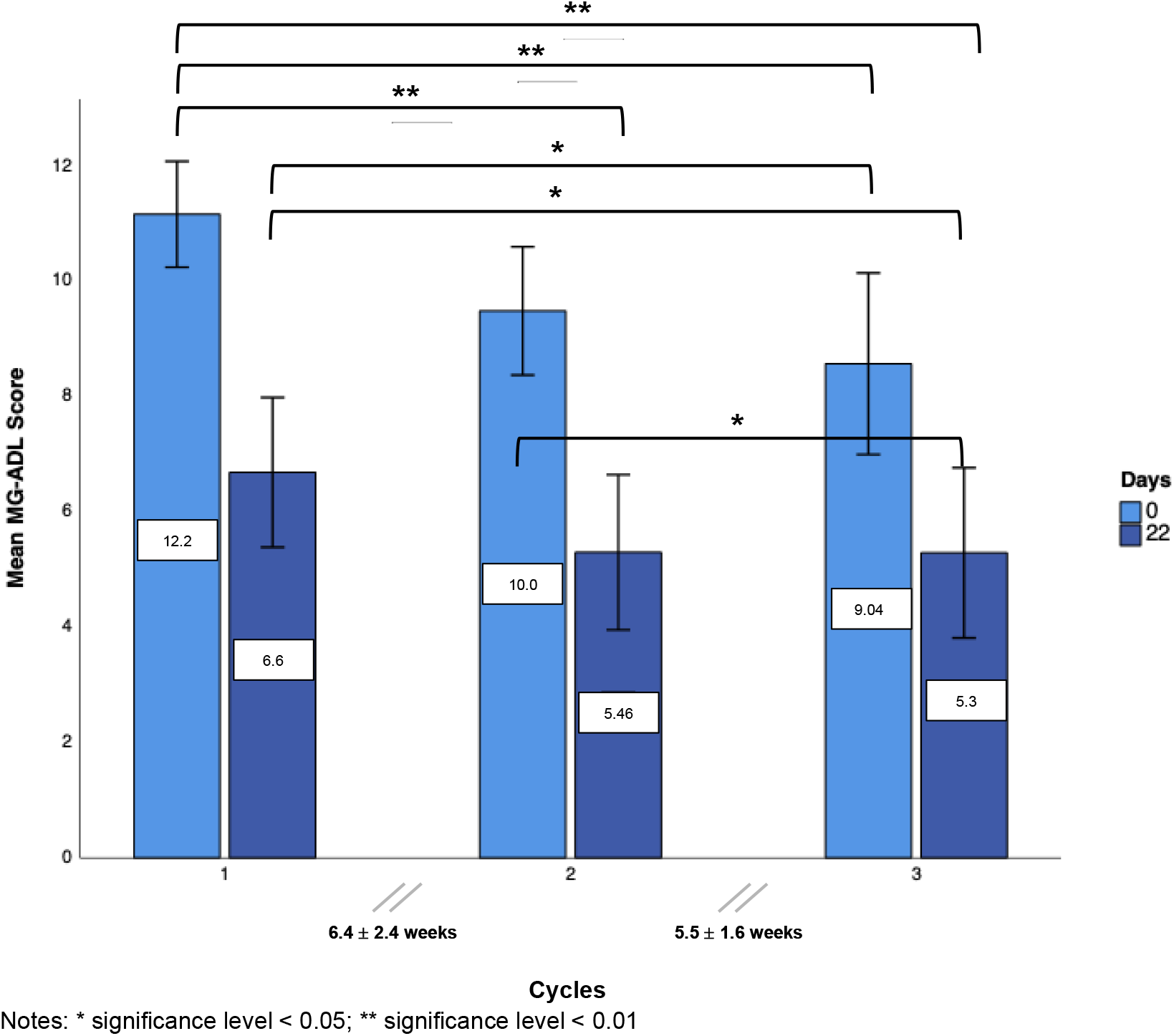
MG-ADL Score variation in the three cycles (N = 25)

The timing of Efgartigimod treatment is bespoke with a varying time between the end of one cycle and the start of the next one depending on patient symptoms. The mean time interval between finishing the first cycle and starting the 2^nd^ cycle was 6.4 weeks (3 – 15.7 weeks, SD 2.4). This interval decreased slightly between the second and third cycles (approximately 5.5 weeks, (3-– 10.9 weeks, SD 1.6) and between the third and fourth cycles (approximately 4.6 (3.0 -6.7) weeks, SD 0.9). In a *post-hoc* analysis, the interval was not statistically significantly correlated to MG-ADL at the beginning of each cycle, and it was not found to be related to an increase in MG-ADL score at the beginning of the next cycle. MG-ADL variation within each cycle (calculated from the difference in MG-ADL at day 22 and day 0) was not correlated with interval duration.

More than a quarter (27.0%, N = 13) of the patients reported a side-effect on the 1^st^ cycle, most of them of mild severity. Three patients reported infections (SARS-CoV-2 infection and urinary tract infection). One patient had severe hypokalemia that required hospitalisation. Seven patients reported flu-like symptoms, one patient reported skin bruising and another reported reduced sensation in the lower legs.

No patients in this study required rescue treatment with IVIg or TPE. Two patients dropped out from this study – one of them did not show any improvement with Efgartigimod after one cycle, and the other patient had a severe adverse reaction (hypokalemia), which was considered to be related to Efgartigimod administration. No deaths were reported.

## DISCUSSION

This is a retrospective real-world study that captured all Efgartigimod-treated patients in the UK from May 2022 to July 2023. The cohort treated were those with long-term refractory MG – the disease duration in the majority of patients was over 10 years, most patients had been on multiple immunosuppressant agents, more than half of patients had received Rituximab, and 54.2% (N = 26) required regular IVIg and/or TPE.

In this group of patients with severe MG, 75.0% were defined as MG responders, as they showed a sustained ≥ 2-point reduction in MG-ADL over 4 weeks, according to what was defined as clinically significant in the ADAPT trial^11^. In our cohort, the mean reductions in MG ADL score were 4.6, 3.9, 3.4 and 4.2 in the first, second, third and fourth cycles, respectively. Though the numbers are small there seemed to be an accumulation of response with average lower baseline scores at the start of Cycle 4 compared to that at the start of cycle 1.

No patients required rescue treatment with IVIG or TPE and no patients had an unplanned admission because of their MG. Efgartigimod had an IVIg and TPE-sparing effect in patients previously dependent on these treatments.

In our population, Efgartigimod seemed to be relatively safe, with about a quarter of patients reporting mild side effects after the first cycle. One patient had a severe metabolic disturbance with hypokalemia which was considered to be related to Efgartigimod, although the physiological explanation for this us unclear and no case reports or drug company notifications exist on this matter.

We observed an excellent response in a few patients (for instance, a patient who started the trial with an MG-ADL score of 11, and, after the 1^st^ cycle, the score lowered to 0, reaching a score of 4 at the beginning of the 2^nd^ cycle, and afterwards keeping a score of 2), but we also detected some cases that seemed to have no response at all (for instance, the one patient that dropped out of the study after the 1^st^ cycle because no difference on the MG-ADL score was observed).

There are some limitations to our study. Although it captured all patients treated with Efgartigimod in the UK between May 22 and July 23, the sample size is small and our average duration of follow-up from the first cycle is 130.9 days (47 – 207, SD 43.2). It was a retrospective real-world study with a heterogeneous group of patients. Our study was also not designed to analyse what factors were associated with response to Efgartigimod. There were no definite criteria for inclusion in the study other than AChR-antibody-positive generalized disease that was not adequately controlled on standard therapies and, depending on access to clinical trials, clinical experience, and access to infusion centre facilities, individual sites may have had different thresholds for patient inclusion. Moreover, the timing of Efgartigimod treatment is variable and dependent on the clinicians’ and patients’ assessment of their disease severity.

The interval between treatments declined after Cycle 1 – likely because the patient and clinician could predict when the symptoms were likely to deteriorate and adjusted the timing of the next cycle to preempt the worsening of symptoms. A significant proportion of patients (at least 35.2%) received home treatments since cycle 2.

We also do not have long-term follow-up data to show whether Efgartigimod has a steroid-sparing effect or not and this is something that should be studied in future.

Our findings are broadly in keeping with those of the ADAPT study^15^, though our patient cohort was slightly different, the average disease duration was longer in our cohort whereas 26% of those in the international trial had not been treated with an oral immunosuppressant agent, all our patients had.

Our findings were also in keeping with data from elsewhere; an Italian centre presented data regarding Efgartigimod efficacy and safety in refractory MG patients^16^, having included 19 patients. After two cycles, a mean 5-point reduction in MG-ADL was observed, minimal manifestation status (MMS) was achieved in 16% of the patients with an improved status was found in 63%^16^.

In our total cohort, a mean 3.9 points reduction in MG-ADL was observed after two cycles. Minimal manifestation state was achieved in 12.5% by the end of the second cycle, and an improved status (considering a 2-point reduction) was obtained in 65.6% of our patients. It is worth noting, however, that both triple negative MG and MuSK patients were also included in the Italian study, whereas in our study only anti-AChR positive patients were included.

Our cohort was mainly composed of patients with refractory long-term MG. As such, our study did not answer the question of where Efgartigimod should fit in the treatment pathway – there can still be potential for its use in situations other than refractory disease. In the UK Efgartigimod is only licensed for AChR-positive MG patients though the ADAPT trial did include MuSK-positive and seronegative patients. There is a rationale for the use of Efgartigimod in these groups, but they have not been studied in detail to date.

## CONCLUSION

Efgartigimod seems to be an efficacious and safe drug for refractory MG patients. However, the profile of the group of patients that are more likely to benefit from Efgartigimod, as well as its exact timing in refractory Myasthenia gravis, remains to be determined. Also, it is not yet clarified if there is an advantage in maintaining other drugs that act in different pathways of the pathophysiology of MG-related neuromuscular junction dysfunction.

Larger prospective studies are also required to establish efficacy and safety in real-world settings.

## Data Availability

All data produced in the present study are available upon reasonable request to the authors.

## ILLUSTRATIONS

### ABBREVIATIONS

IQR: interquartile range
IVIg: intravenous immunoglobulin g
MG: generalised MG
MG: Myasthenia Gravis
MG-ADL: Myasthenia Gravis Activities of Daily Living
NSIST: non-steroid immunosuppression treatment
TPE: plasmapheresis

## SUPPLEMENTARY MATERIAL

**Table 1.** MG-ADL variation in the three first cycles (N = 25)

| (I) Time | (J) Time | Mean<br>Difference (I-J) | Std. Error | Sig.<sup>b</sup> | 95% Confidence Interval for<br>Difference<sup>b</sup> |  |
| --- | --- | --- | --- | --- | --- | --- |
|  |  |  |  |  | Lower Bound | Upper Bound |
| Cycle 1, Day 0 (C1D0) | C1D22 | 5.542* | .697 | <.001 | 3.261 | 7.823 |
|  | C2D0 | 2.125 | .806 | .221 | -.514 | 4.764 |
|  | C2D22 | 6.708* | .688 | <.001 | 4.457 | 8.959 |
|  | C3D0 | 3.125* | .904 | .032 | .167 | 6.083 |
|  | C3D22 | 6.875* | .753 | <.001 | 4.411 | 9.339 |
| Cycle 1, Day 22 (C1D22) | C1D0 | -5.542* | .697 | <.001 | -7.823 | -3.261 |
|  | C2D0 | -3.417* | .857 | .009 | -6.224 | -.610 |
|  | C2D22 | 1.167 | .693 | 1.000 | -1.103 | 3.436 |
|  | C3D0 | -2.417 | .895 | .191 | -5.346 | .512 |
|  | C3D22 | 1.333 | .846 | 1.000 | -1.437 | 4.104 |
| Cycle 2, Day 0 (C2D0) | C1D0 | -2.125 | .806 | .221 | -4.764 | .514 |
|  | C1D22 | 3.417* | .857 | .009 | .610 | 6.224 |
|  | C2D22 | 4.583* | .796 | <.001 | 1.977 | 7.190 |
|  | C3D0 | 1.000 | .493 | .812 | -.613 | 2.613 |
|  | C3D22 | 4.750* | .854 | <.001 | 1.954 | 7.546 |
| Cycle 2, Day 22 (C2D22) | C1D0 | -6.708* | .688 | <.001 | -8.959 | -4.457 |
|  | C1D22 | -1.167 | .693 | 1.000 | -3.436 | 1.103 |
|  | C2D0 | -4.583* | .796 | <.001 | -7.190 | -1.977 |
|  | C3D0 | -3.583* | .819 | .003 | -6.263 | -.904 |
|  | C3D22 | .167 | .527 | 1.000 | -1.559 | 1.892 |
| Cycle 3, Day 0 (C3D0) | C1D0 | -3.125* | .904 | .032 | -6.083 | -.167 |
|  | C1D22 | 2.417 | .895 | .191 | -.512 | 5.346 |
|  | C2D0 | -1.000 | .493 | .812 | -2.613 | .613 |
|  | C2D22 | 3.583* | .819 | .003 | .904 | 6.263 |
|  | C3D22 | 3.750* | .740 | <.001 | 1.326 | 6.174 |
| Cycle 3, Day 22 (C3D22) | C1D0 | -6.875* | .753 | <.001 | -9.339 | -4.411 |
|  | C1D22 | -1.333 | .846 | 1.000 | -4.104 | 1.437 |
|  | C2D0 | -4.750* | .854 | <.001 | -7.546 | -1.954 |
|  | C2D22 | -.167 | .527 | 1.000 | -1.892 | 1.559 |
|  | C3D0 | -3.750* | .740 | <.001 | -6.174 | -1.326 |

Based on estimated marginal means
\*. The mean difference is significant at the .05 level.
b. Adjustment for multiple comparisons: Bonferroni.

